# Benefit of targeted sampling for lymphatic filariasis surveillance in Samoa depends on antigen prevalence

**DOI:** 10.1101/2024.10.11.24315286

**Authors:** Helen J Mayfield, Benn Sartorius, Angus McLure, Stephanie J Curtis, Beatris Mario Martin, Sarah Sheridan, Robert Thomsen, Rossana Tofaeono-Pifeleti, Satupaitea Viali, Patricia M Graves, Colleen L Lau

## Abstract

**Background:** In Samoa, lymphatic filariasis (LF) remains endemic. Targeted sampling strategies based on locations of known infections could be more efficient than random sampling for locating infected individuals and hotspots, providing valuable information to develop more efficient and cost-effective interventions. However, the level of benefit may depend on the prevalence of the chosen indicator in the area being surveyed. This study aims to assess the efficiency of targeted versus random sampling for identifying LF antigen (Ag)- and microfilaria (Mf)-positive individuals in Samoa for varying background Ag prevalence levels.

**Methodology:** In 2023, six primary sampling units (PSUs) were surveyed using random and targeted sampling strategies. PSUs were selected based on Ag prevalence in 2019, including two low (3-5%), medium (6-7%) and high Ag prevalence (13-17%). The randomly selected group included residents aged ≥5 years in 15 houses per PSU. The targeted group included residents aged ≥5 years in up to eight households within 200 metres of a household where Ag-positive resident(s) were identified in 2019. Finger prick blood samples were tested for Ag and Ag-positive samples were examined for microfilaria (Mf).

**Principal Findings:** The targeted sampling strategy (n=400 people) identified more positives (57 Ag-positive, 23 Mf-positive) than the random sampling strategy (n=494, 39 Ag-positive, 16 Mf-positive), with an overall targeted:random sampled case ratio of 1.8 (95% CI 1.3-2.5) for Ag and 1.8 (95% CI 1.1-3.1) for Mf. Gain in efficiency was greatest in medium prevalence PSUs for both Ag-positives (ratio=2.4, 95% CI 1.3-5.2) and Mf-positives (ratio=2.6, 95% CI 0.9-12.8).

**Conclusions:** In Samoa, a targeted sampling strategy was more efficient for locating Ag-positive and Mf-positive individuals compared to random sampling, with the highest efficiency gain in medium Ag prevalence settings. The findings have design implications for LF surveillance in Samoa and other Pacific Island countries.

**Author Summary:** Effective surveillance activities are essential for achieving elimination targets for lymphatic filariasis (LF). Cost-effective surveillance strategies are needed to locate infections for targeted treatment of individuals or high-risk communities. The aim of this study was to investigate the efficiency of targeted sampling compared to random sampling for locating antigen (Ag)-positive and microfilaria (Mf)-positive individuals in Samoa and explore how differences in efficiency gains depend on the Ag prevalence in the village. In 2023, six villages with varying Ag prevalence in 2019 were surveyed using both random and targeted sampling strategies. For the random group, we selected 15 houses in each village and invited all household residents aged five years and over to participate. The targeted group consisted of residents in these same villages who lived within 200 metres of an Ag-positive participant from the 2019 survey. Finger prick blood samples were collected and tested for Ag and Mf.

Overall, significantly more cases were identified per person tested by using the targeted strategy. This gain was particularly evident in the medium prevalence villages. These results show that targeted surveys of households neighbouring the residence of an Ag-positive person may be more efficient than random sampling, particularly in medium Ag prevalence villages. The findings have specific design implications for future LF surveys in Samoa and other Pacific Island countries.

## Introduction

Effective monitoring and surveillance activities are essential for achieving elimination targets for neglected tropical diseases (NTDs). For programs such as the Global Program to Eliminate Lymphatic Filariasis (GPELF) [1] to successfully achieve their goals, robust surveillance data are needed to inform decisions on interventions such as mass drug administration (MDA) [2, 3], or locate hotspots of residual infection that could hamper elimination efforts [4, 5]. In a pre-elimination context where the disease is still endemic, detecting ongoing transmission or potential resurgence remains a priority [6].

When designing surveillance activities, there are various potential sampling strategies for selecting who and where to sample. These include randomly selecting participants or households, or targeted surveys of sub-populations, demographic groups, or locations [7]. For surveys where the objective is to estimate overall prevalence of an infection marker, random sampling is most appropriate to obtain a population representative sample. For example, the WHO-recommended Transmission Assessment Survey (TAS) for lymphatic filariasis (LF) is a population representative survey (typically of young children) designed to provide an estimate of Ag prevalence in that age group. [6].

Another potential surveillance objective is to cost-effectively locate infected people or sub-populations for targeted response; for this situation, random sampling of the population may not be the best course of action. For LF, targeting known high prevalence groups such as adult males or residents of known hotspots may locate more infections per person sampled [2, 8]. Various data-driven approaches have been developed for guiding targeted sampling, including data modelling [9] and machine learning approaches [4]. These strategies are generally data-intensive and require expertise and resources that are often not available to program managers or decision makers in LF-endemic countries.

Another option for targeted surveillance is to sample participants based on their proximity to or contact with known infected people, an approach variously referred to as contact tracing, reactive case finding, or snowball sampling. In the case of LF, infection would most likely refer to the standard infection indicators, antigen (Ag) and microfilaria (Mf). The strategy used to direct targeted sampling can be based on people living in the same or nearby households, or frequently attending the same locations such as school, work, place of worship or other community gatherings. The efficiency of targeted sampling strategies depends on several factors including the way that index cases were identified, the number of targeted samples for each index case [10], and the overall prevalence of the chosen indicator [4].

In Samoa, Ag prevalence is highly heterogenous between villages [2, 11] and multiple rounds of MDA have not been sufficient to break transmission [2]. A national survey in 2018 of 35 primary sampling units (PSUs) estimated Ag prevalence at the PSU level in participants aged ≥5 years ranging from 0% (1-sided 97.5% CI 0-3.7%) to 10.3% (95% CI 5.9-17.6%), and demonstrated that Ag-positive persons were geographically clustered at the household and village levels [2]. This study aims to investigate the efficiency of targeted sampling compared to random sampling for locating Ag-positive and Mf-positive people in Samoa in 2023. Specifically, the objectives of this study were to i) evaluate if targeted sampling of near neighbours (within 200 m) of Ag-positive individuals was more efficient for locating additional Ag-positive or Mf-positive persons compared to random sampling; and ii) determine if there is an Ag prevalence threshold or range where targeted sampling of near neighbours is more efficient than random sampling.

## Methods

### Study area

Samoa is located in the South Pacific and has two main islands, Upolu and Savai’i, with a total population of approximately 200,000 residents [12]. The wet tropical climate creates year-round suitable conditions for LF vector species, primarily *Aedes polynesiensis* [13]. Samoa has a long history of LF and has carried out many rounds of MDA since 1965 as part of ongoing elimination efforts [14]. In 2018, Samoa was the first country to implement nation-wide triple-drug MDA for LF using ivermectin, diethylcarbamazine and albendazole [15]. A second triple-drug MDA round was scheduled for 2019, but this was delayed until September 2023 because of public health emergencies. While there are currently no routine surveillance programs for LF in Samoa, operational research surveys conducted in 2018 (1.5 to 3.5 months post-MDA) and 2019 (six to eight months post-MDA) in 35 primary sampling units (PSUs) provided detailed epidemiological data on Ag and Mf prevalence [2, 16].

### Ethics approval and informed consent

Ethics approvals were obtained from the Samoa Ministry of Health and The University of Queensland Human Research Ethics Committee (protocol 2021/HE000895). The study was conducted in close collaboration with the Samoa Ministry of Health, the World Health Organization (WHO) country office in Samoa, and the Samoa Red Cross. Prior to entering a village, permission was granted from village leaders to conduct the study. Verbal and written informed consent were obtained for all participants, or from the parents or guardians of participants who were less than 18 years old.

### Survey design

In 2023, eight PSUs were selected from the 35 PSUs included in the 2018 and 2019 surveys [16]. To compare the efficiency benefits of targeted sampling in different transmission settings, PSUs sampled in 2023 were selected based on Ag prevalence in the 2019 survey. Two PSUs each with low (3-5%), medium (6-7%) and high (13-17%) Ag prevalence were surveyed using both random and targeted sampling strategies. Two PSUs with no Ag-positive participants identified in 2019 were also surveyed but were not included in this analysis as they did not include any known Ag-positive households to guide the targeted survey. For logistical reasons, all selected PSUs in 2023 were in Upolu (Fig. 1). The survey was conducted in February-March 2023, approximately 4.5 years after the first round of triple-drug MDA in August 2018, and prior to the second round of triple-drug MDA in September 2023.

**Fig. 1.**
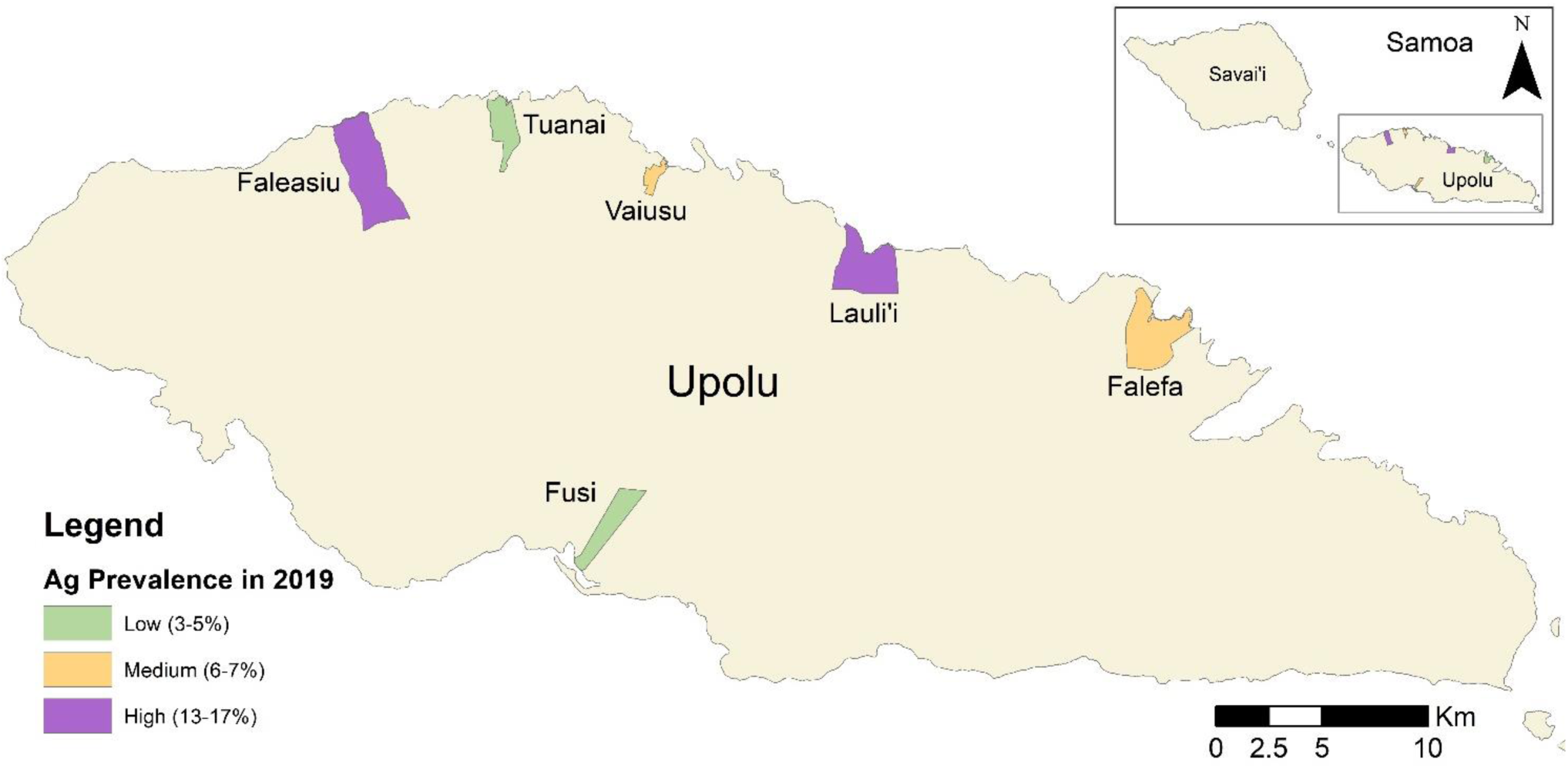
Location of the six primary sampling units (PSUs) in Samoa that were surveyed for lymphatic filariasis in 2023 using both random and targeted sampling stratergies

Each PSU included randomly selected and targeted sampling survey components. For the randomly selected component, 15 households were chosen based on a virtual walk method (previously described) that provided a spatially representative sample of households in the PSU [2]. If less than 60 participants aged ≥10 years were enrolled in a PSU after 15 houses were surveyed, houses neighbouring those of existing participants were included until the sample size was met.

For the targeted sampling approach, households of Ag-positive participants (index cases) from the 2019 survey [16] were used as seed households if it was confirmed that the index case had resided in the house for any time since the 2019 survey.

A buffer of 200 m radius (selected based on density of households and the estimated 150 m flight range of the primary vector) was defined around each household. For each seed household, up to eight households within this buffer were included for targeted sampling, starting with the closest household and moving outwards. To maintain privacy, targeted households were not informed of the household location or identity of the Ag-positive index case from the seed household. If a household selected for targeted sampling was also sampled as part of the randomly selected group, it was included in the analysis for both groups. Members of seed households (including the index case) were tested but were not included in the targeted group.

All residents of selected households aged ≥5 years were invited to participate. If a household member was eligible to participate but absent at the time of the visit, the field team returned to the house that evening or the next day where feasible. Consistent with protocols described elsewhere [2], consenting participants were asked questions on their demographics, and household GPS coordinates were collected using a smart phone. Household surveys were conducted primarily by bilingual (English and Samoan) fieldworkers.

### Testing for antigen and microfilaria

For each participant, finger prick blood samples (400uL) were collected in heparin microtainers and tested for Ag using Abbott Alere Filariasis Test Strips (FTS) (Scarborough, ME, USA). FTS were read at ten minutes according to manufacturer’s instructions. For Ag-positive samples, three thick blood smears (slides) were prepared according to WHO guidelines [6], with three 20μL lines of blood per slide, i.e. total 60 μL. Slides were dried for 72 hours and then dehaemoglobinised in water for 10 to 15 minutes before being dried, fixed with methanol and stained. Slides were read by a trained technician in the field laboratory using a microscope, and a sample was considered Mf-positive if any Mf were observed on either slide. All Mf-positive participants were informed of their results and offered treatment with the same drugs and dosage used during the 2018 triple-drug MDA.

### Statistical Analysis

For both Ag and Mf, we calculated the prevalence of positive individuals for randomly selected and targeted groups. Values were calculated for overall Ag prevalence and for each of the three 2019 Ag prevalence categories (low, medium and high) and by PSU. All data were analysed using Stata Version 18.0 [17]. We calibrated the sampling weights using the “survwgt” command to standardise to the census population by PSU. The weighted prevalence was estimated using the ‘svyset’ command from the ‘svy’ package to account for the sampling design and probability of selection. The proportion of Ag-positive and Mf-positive households (those with at least one positive resident) was calculated for each group, adjusting for the household level probability of selection. All estimates were computed with 95% jackknife confidence intervals (CIs)

To evaluate the efficiency of targeted compared to random sampling in terms of Ag-positive and Mf-positive individuals found per person sampled, we performed a comparative analysis of the absolute number of positive cases identified through each approach. Given that the denominators (number of people sampled) for the targeted and randomly selected groups were different, we multiplied the proportion of positives for the targeted sample with the denominator attained for the random sample. We then estimated the ratios of absolute counts of positive cases obtained from targeted versus random sampling (overall, by prevalence setting and PSU) using a Poisson distribution with 95% confidence intervals. This was done to determine if the targeted sampling strategy yielded a significantly higher number of Ag- and Mf-positive cases compared to the random sampling strategy.

## Results

### Participants

A total of 899 participants were enrolled from 190 households across the six PSUs. By chance, five (3%) households, with a combined total of 18 participants, were included in both the randomly selected and targeted groups. Valid FTS results were obtained from 876 (97.4%) of enrolled participants in 186 households (97.9%). Age and sex of participants were similar across both groups (Table 1).

**Table 1.** Number of participants with valid antigen (Ag) test results stratified by sex and age for randomly selected and targeted groups in six primary sampling units (PSUs) in Samoa, 2023.

|  | 2019 Ag prevalence category |  | Randomly selected group | Targeted group |
| --- | --- | --- | --- | --- |
| Households n (%) | Total |  | 92 | 98 |
|  | Low |  | 30 (32.6%) | 26 (26.5%) |
|  | Medium |  | 31 (33.7%) | 31 (31.6%) |
|  | High |  | 31 (33.7.1%) | 41 (41.8%) |
| Participants n (%) | Total |  | 494 | 400 |
|  | Low |  | 163 (33.0%) | 97 (24.3%) |
|  | Medium |  | 159 (32.2%) | 141 (35.3%) |
|  | High |  | 172 (34.8 %) | 162 (40.5%) |
| Sex n (%) | Total | Male | 224 (45.3%) | 179 (44.8%) |
|  |  | Female | 270 (54.7%) | 221 (55.2%) |
|  | Low | Male | 82 (50.3%) | 51 (52.6%) |
|  |  | Female | 81 (49.7%) | 46 (47.4%) |
|  | Medium | Male | 67 (42.1%) | 63 (44.7%) |
|  |  | Female | 92 (57.9%) | 78 (55.3%) |
|  | High | Male | 75 (43.6%) | 65 (40.1%) |
|  |  | Female | 97 (56.4%) | 97 (59.9%) |
| Age in years mean (range) | Total |  | 22 (5-93) | 24 (5-86) |
|  | Low |  | 20 (5-93) | 23 (5-79) |
|  | Medium |  | 18 (5-78) | 22 (5-76) |
|  | High |  | 26 (5-87) | 27 (5-86) |

Twenty house locations of Ag-positive participants from the 2019 survey were confirmed and used as seed households for the 2023 study. The mean distance between a seed and a targeted sampled household was 77 m (range: 12m - 191m) (Table 2).

**Table 2.** Number of seed households, number of targeted households per seed, and distance between the seed and targeted household for each 2019 Ag prevalence category (low [3-5%], medium [6-7%] and high [13-17%]).

| **2019**<br>**Ag prevalence**<br>**category** | **Ag-positive seed**<br>**households (n)** | **Number of targeted**<br>**households per seed,**<br>**mean (range)** | **Distance between seed and**<br>**targeted households in**<br>**metres, mean (range)** |
| --- | --- | --- | --- |
| Total | 20 | 4.9 (1-8) | 77 (12-191) |
| Low | 4 | 6.5 (6-7) | 69 (13-186) |
| Medium | 5 | 6.2 (2-7) | 93 (23-175) |
| High | 11 | 3.7 (1-8) | 71 (12-191) |

### Antigen prevalence

Ag-positive residents were found in both the randomly selected and targeted groups, and in all six PSUs (Fig 2a). In the randomly selected group, observed Ag prevalence in 2023 for the two PSUs in the low Ag prevalence remained within the range for this category (Supplementary S1 Fig, Supplementary S1 Table). In Lauli’i (selected in the high prevalence category) the observed prevalence in 2023 of 9.8% (95% CIs 4.2-20.9%) was closer to the range of the medium category. In Vaiusu (medium prevalence category PSU) the observed Ag prevalence in 2023 of 2.4% (95% CIs 0.7-8.7%) was within the range of the low prevalence category.

**Fig 2.**
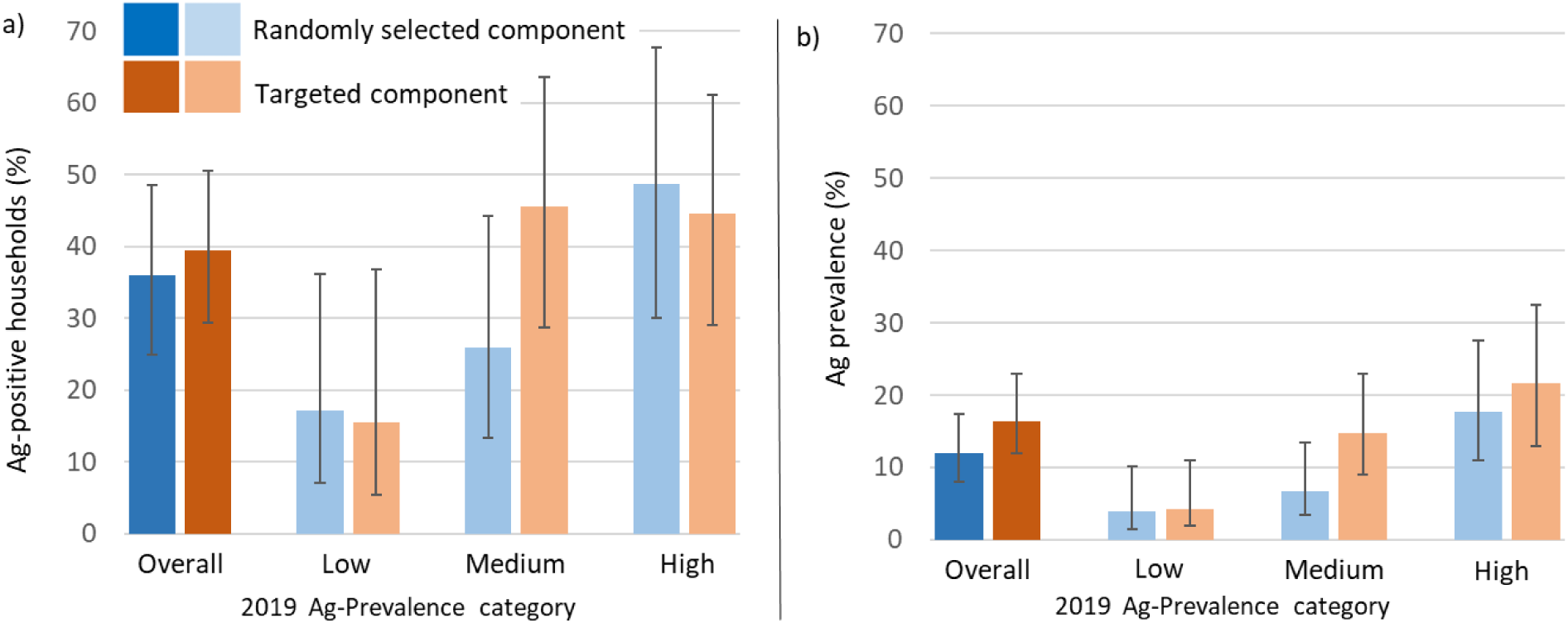
a) Percentage of Ag-positive households and b) adjusted Ag prevalence and 95% confidence intervals in the randomly selected (blue) and targeted (orange) groups for each 2019 Ag prevalence category (low [3-5%], medium [6-7%] and high [13-17%]) in six primary sampling units (PSUs) surveyed in 2023 in Samoa.

The proportion of surveyed households with at least one Ag-positive resident was not significantly different in the targeted group (39.4% of 98 households, 95% CI: 29.3-50.6%) compared to the randomly selected group (35.9% of 92 households, 95% CIs 24.9-48.6%) (Table 3). Fig 2a shows that the largest difference between the two groups was in the category with medium Ag prevalence in 2019, with 45.6% (95% CI: 28.7-63.5%) of households in the targeted group and 25.9% (95% CI: 13.30-44.20%) in the randomly selected group having at least one Ag-positive participant in 2023, although this difference was not statistically significant.

Overall adjusted Ag prevalence was higher but not statistically different in the targeted group (17.0%, 95% CI: 12.0-24.6%) compared to the randomly selected group (11.9%, 95% CI: 8.0-17.4%). This trend was consistent in each of the 2019 Ag prevalence categories, although the magnitude of the difference varied (Fig 2b, Supplementary S2 Table).

### Microfilaria prevalence

Households with Mf-positive residents were found in both the randomly selected and targeted groups, and in all six PSUs (Fig 3a). Overall, the proportion of households in the targeted group that had at least one Mf-positive resident (18.7% of 98, 95% CI: 11.5-28.8%) was not significantly different to the randomly selected group (22.5% of 92, 95% CI: 13.2-35.6%). The overall adjusted Mf prevalence was not significantly higher in the targeted group (7.4%, 95% CI: 3.7-14.3%) compared to the randomly selected group (6.1%, 95% CIs 3.7- 9.9%), as shown in Fig 3b and Supplementary S3 Table.

**Fig 3.**
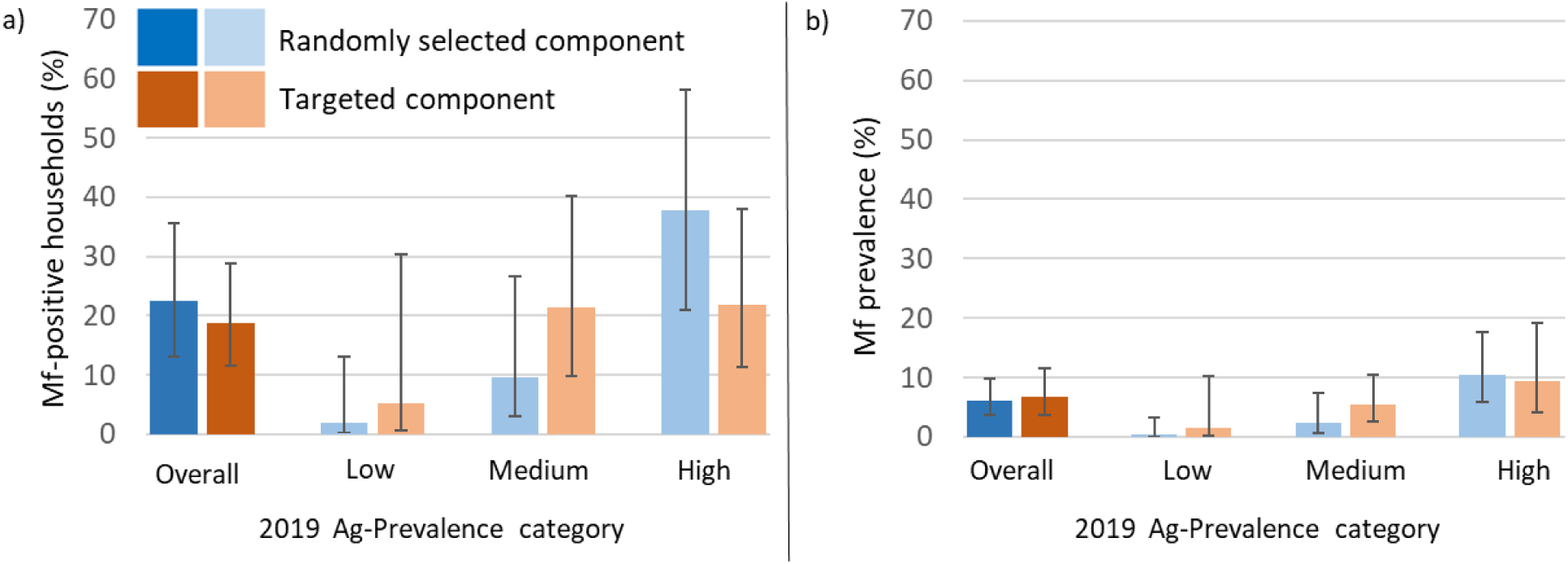
a) Percentage of Mf-positive households and b) adjusted Mf prevalence and 95% confidence intervals in the randomly selected (blue) and targeted (orange) groups for each 2019 Ag prevalence category (low [3-5%], medium [6-7%] and high [13-16%]) in six primary sampling units (PSUs) surveyed in 2023 in Samoa.

### Gains in sampling efficiency

Overall, the ratio of positive cases identified in the targeted group versus the randomly selected group was significantly higher at 1.81 (95% CI 1.32-2.55) for Ag and 1.78 (95% CI 1.10-3.11) for Mf, respectively. There was a higher ratio (i.e., gain in efficiency) in medium prevalence PSUs for Ag-positives (2.4, 95% CI 1.3-5.2) and for Mf-positives (2.6, 95% CI 0.9-12.8) (Fig 4, Supplementary Table S4). There was no significant difference in this ratio for the low prevalence setting for either Ag or Mf. In high prevalence setting there was a significant gain in Ag-positive case yield for the targeted sampling strategy (ratio=1.51, 95% CI 1.02-2.36), but not for Mf-positives.

**Fig 4.**
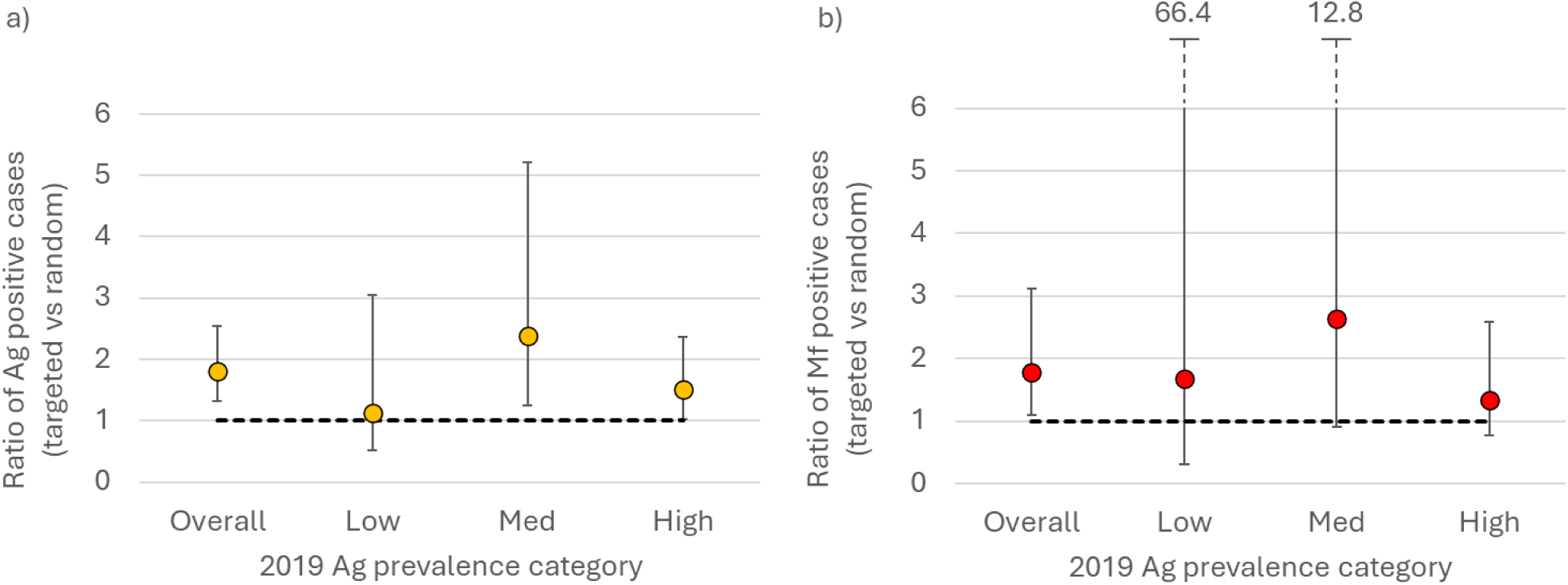
Ratio of a) Ag-positive individuals and b) Mf-positive individuals in the targeted group compared to the randomly selected group, including 95% confidence intervals. Values presented for each 2019 Ag prevalence category (low [3-5%], medium [6-7%] and high [13-17%]) - in six primary sampling units (PSUs) surveyed in 2023 in Samoa.

## Discussion

Our study found that targeted sampling based on proximity to the household of an Ag-positive person was significantly more efficient for locating additional Ag-positive and Mf-positive people in Samoa. Overall, targeted sampling identified almost double the number of positive cases compared to random sampling. The efficiency gain was particularly evident in the medium Ag prevalence villages (6-7%), with over 2.4 and 2.6 Ag-positive and Mf-positive cases identified in the targeted group for every positive case identified in the randomly selected group, respectively. These results provide evidence that targeted sampling of near neighbours of Ag-positive people could provide an efficient strategy for elimination programs to identify and target local transmission foci for treatment or more intensive surveillance. From an operational perspective, a gain in sampling efficiency equates to fewer resources required to find and treat more infections. The benefits, however, need to be matched with other on-the-ground considerations, such as maintaining the anonymity of infected people, especially in close-knit villages and communities.

The findings presented in this study support previous work by Gass *et. al.* in Ethiopia and Tanzania showing that different sampling strategies are preferable depending on the background prevalence [18]. We found targeted sampling to be no more efficient than random sampling in the low Ag prevalence settings, supporting findings from simulation studies which found that random sampling was more efficient than targeted (snowball) sampling at locating LF hotspots in settings with a low (1%) Ag prevalence [10]. Although a small but significant gain in efficiency was observed in the high prevalence PSUs, this was likely due to the observed Ag prevalence of one the high prevalence PSUs in 2019 (Lauli’i) falling within our definition of medium prevalence by 2023. An increased sample size is needed to confirm this finding as well as further research into prevalence ranges where targeted sampling would be more efficient.

The influence of the background Ag prevalence on sampling efficiency can be considered in terms of the potential gain for a given scenario. In very low prevalence settings, the low number of overall infections means that there is less potential for absolute gain in the number of infections located, regardless of sampling strategy. In relative terms however, even small gains in efficiency will be beneficial in countries nearing elimination. At very high prevalence, such as that seen in Faleasiu, it is likely that a relatively large proportion of the randomly selected group live within the pre-determined buffer distance of an infected household. This results in little if any difference in the distance to the nearest Ag-positive household between the randomly selected and targeted groups, and therefore little or no gain is seen from a targeted sampling approach in this instance. Further work remains to refine the prevalence range, cut-offs or other scenarios where targeted sampling based on household location would be more efficient than random sampling, acknowledging that these may differ between settings.

The efficiency gains of a targeted sampling strategy based on spatial proximity to known infections are a result of the clustering of infections. In the context of LF elimination, this pattern may be influenced by several factors, including environmental factors [19] and the coverage of national or regional interventions such as MDA. Currently, there is limited data on the effect of widescale MDA on clustering of infections. Targeted sampling based on the location of infected people also relies on finding an initial index case or seed household. For areas that have not been recently surveyed, modelling and predictive mapping can potentially provide useful guidance on where to look [10, 20–22]. Other opportunities for identifying index cases include transmission assessment surveys (TAS) that form part of the process for validating elimination [23, 24], opportunistic screening, or clinical presentations of LF.

Our results should be considered in light of the study’s limitations. The study design was not powered to detect significant differences in prevalence between groups at the levels observed in the low and high prevalence categories. In addition, there was a 4.5-year gap between identifying the index cases in 2019 and surveying the targeted households in 2023, with only 11 of the 20 positive seed households still having confirmed Ag-positive resident(s) at the time of this study. A shorter delay between identifying the seed house and testing the neighbouring households would more closely resemble a real-time targeted surveillance strategy and potentially further improve efficiency.

The findings presented here have specific design implications for future LF surveys in Samoa and other similar Pacific Island countries. Better understanding of the potential gains from applying different sampling strategies can help to optimise resource use and improve the information available to decision makers. The results demonstrate that the expected prevalence should be considered when designing surveys for monitoring and surveillance to support LF elimination. In particular, targeted surveys of households neighbouring the residence of an Ag-positive index case may be most efficient within a certain Ag prevalence range. In medium prevalence settings, targeted sampling of nearby households may prove to be better than random sampling strategies, and it may be effective for further narrowing down transmission foci in high burden contexts, facilitating a more efficient resource deployment. In high prevalence settings, targeted sampling of near neighbours offers little benefit over random sampling and blanket treatment approach is likely a more efficient use of resources.

## Data Availability

Data used in this paper were collected during field surveys in Samoa. Communities in Samoa are small (some with less than 200 inhabitants) and sharing individual level data could enable identification of individual participants, and violating the conditions of the study’s ethics approval. For requests relating to data access, please contact the Human Ethics Department at the University of Queensland citing protocol 2021/HE000895—. if you would like access to the data. All relevant data at the primary sampling unit level has been included in the supplementary material.

## Acknowledgements

We are extremely grateful to our colleagues at the Samoa Red Cross for their continued and invaluable support as field teams during SaMELFS surveys, as well as staff from the Samoa Ministry of Health. We would also like to acknowledge the significant in-country support provided by Lepaitai Hansell and Dyxon Hansell at the WHO office in Apia and extend our sincere thanks for their assistance during the survey preparation and fieldwork. We gratefully acknowledge Shannon Hedtke for reading the Mf slides from the 2018 and 2019 SaMELFS surveys, and Jane Sinclair, Ramona Muttucumaru Maddison Howlett and Jessica Scott for their assistance with fieldwork for the 2023 survey. Tara Brant, Ula Mageo, Lynette Suiaunoa-Scanlan, Emily Dodd and Kimberly Won were instrumental in securing Filarial Test Strips (FTS) for the 2023 field work. Finally, we would like to acknowledge Filipina Amosa-Lei Sam for her assistance with translating research documents from English to Samoan.

## Data Sharing

Data used in this paper were collected during field surveys in Samoa. Communities in Samoa are small (some with less than 200 inhabitants) and sharing individual level data could enable identification of individual participants, and violating the conditions of the study’s ethics approval. For requests relating to data access, please contact the Human Ethics Department at the University of Queensland citing protocol 2021/HE000895”. if you would like access to the data. All relevant data at the primary sampling unit level has been included in the supplementary material.

## Supplementary

**Supplementary S1 Fig.**
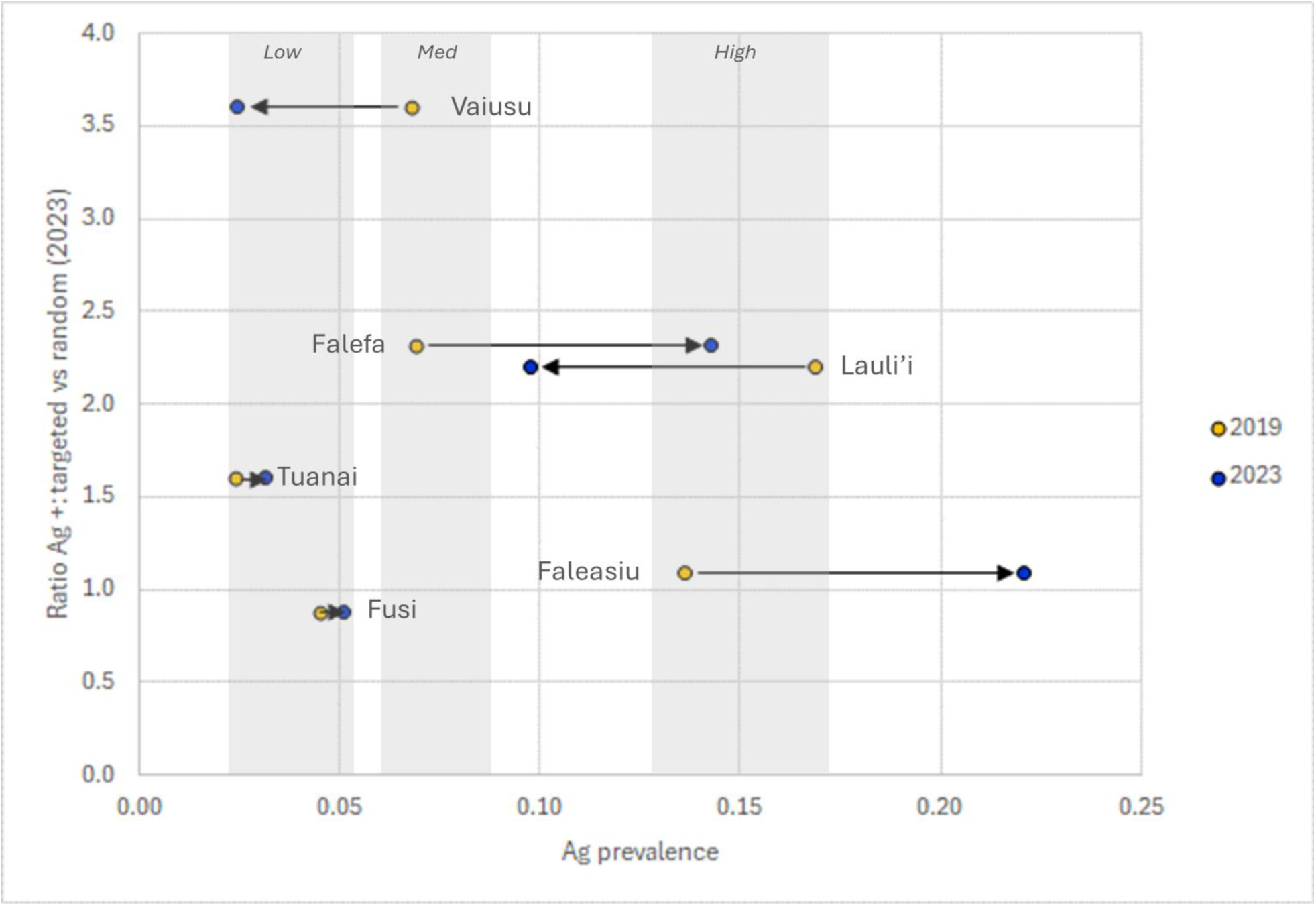
Ratio of antigen-positive individuals to microfilaria-positive individuals by PSU Ag prevalence in Samoa (2019 and 2023 Ag prevalence shown). Shading represents the cut-offs for the low, medium and high 2019 Ag prevalence categories used in this analysis. Values for 2023 are for the randomly selected group.

**Supplementary S1 Table.** Raw and adjusted Ag-prevalence (with 95% confidence intervals) in the targeted and randomly selected groups for each PSU in the 2019 Ag prevalence categories - low (3-5%), medium (6-7%) and high (13-17%) - in six primary sampling units (PSUs) in Samoa in 2023. Ratio of Ag-positive participants in the targeted vs random groups is also shown.

| Ag prev cat (2019) | PSU | 2019 Ag prevalence (95% CI)* | Targeted group |  |  |  |  | Random group |  |  |  | Ratio |
| --- | --- | --- | --- | --- | --- | --- | --- | --- | --- | --- | --- | --- |
|  |  |  | Ag-neg | Ag-pos | Ag-pos (rescaled**) | Total | 2023 Ag prevalence (95% CI) | Ag-neg | Ag-pos | Total | 2023 Ag prevalence (95% CI) |  |
| Low | Fusi | 4.5<br>(1.2-11.0) | 50 | 2 | 3.5 | 52 | 3.9<br>(0.9, 15.3) | 87 | 4 | 91 | 5.5<br>(1.8, 19.0) | 0.9<br>(0.3-3.2) |
|  | Tuanai | 2.4<br>(0.4-7.0) | 43 | 2 | 3.2 | 45 | 4.4<br>(1.2, 15.7) | 70 | 2 | 72 | 3.1<br>(0.6, 13.9) | 1.6<br>(0.4-13.2) |
| Medium | Vaiusu | 6.8<br>(2.7-13.5) | 91 | 7 | 7.2 | 98 | 7.1<br>(3.1, 15.7) | 99 | 2 | 101 | 2.5<br>(0.7, 8.7) | 3.6<br>(1-29.8) |
|  | Falefa | 6.9<br>(2.3-14.9) | 31 | 12 | 16.2 | 43 | 27.9<br>(15.9, 44.1) | 51 | 7 | 58 | 14.3<br>(6.2, 29.4) | 2.3<br>(1.1-5.8) |
| High | Faleasiu | 13.6<br>(7.8-21.2) | 58 | 17 | 16.3 | 75 | 22.7<br>(12.4, 37.9) | 57 | 15 | 72 | 22.1<br>(12.6, 35.7) | 1.1<br>(0.7-1.9) |
|  | Laulii | 16.9<br>(9.1-27.2) | 69 | 17 | 19.8 | 86 | 19.8<br>(8.9, 38.3) | 91 | 9 | 100 | 9.8<br>(4.2, 20.9) | 2.2<br>(1.2-4.8) |

\* Taken from Mayfield, H.J., B. Sartorius, S. Sheridan, M. Howlett, B.M. Martin, R. Thomsen, R. Tofaeono-Pifeleti, S. Viali, P.M. Graves, and C.L. Lau, Ongoing transmission of lymphatic filariasis in Samoa 4.5 years after one round of triple-drug mass drug administration. PLOS Neglected Tropical Diseases, 2024. 18(6): p. e0012236.
\*\* Rescaled to match the denominator in the random sample

**Supplementary S2 Table.** Percentage of Ag-positive households and adjusted Ag prevalence and 95% confidence intervals in the targeted and randomly selected groups for each 2019 Ag prevalence category - low (3-5%), medium (6-7%) and high (13-17%) - in six primary sampling units (PSUs) in Samoa in 2023.

| Group | 2019 Ag prevalence category | Households (n) | Percent Ag-positive households (95% CIs) | Participants (n) | Survey-weighted Ag? prevalence (95% CIs) |
| --- | --- | --- | --- | --- | --- |
| Targeted | Overall | 98 | 39.4% (29.3-50.6%) | 399 | 16.4% (11.7-22.5%) |
|  | Low | 26 | 15.4% (5.4-36.8%) | 97 | 4.2% (2.0-11.0%) |
|  | Medium | 31 | 45.6% (28.7-63.5%) | 141 | 14.8% (9.2-22.9%) |
|  | High | 41 | 44.5 (29.1-61.1%) | 161 | 21.6% (13.7-32.5%) |
| Randomly selected | Overall | 92 | 35.9% (24.9-48.6%) | 494 | 11.9% (8.0-17.4%) |
|  | Low | 30 | 17.1% (7.1-36.1%) | 163 | 3.9% (1.4-10.1%) |
|  | Medium | 31 | 25.9 (13.3-44.2%) | 159 | 6.8% (3.4%-13.4%) |
|  | High | 31 | 48.7% (30.1-67.7%) | 172 | 17.8% (11.0-27.6%) |

**Supplementary S3 Table.**
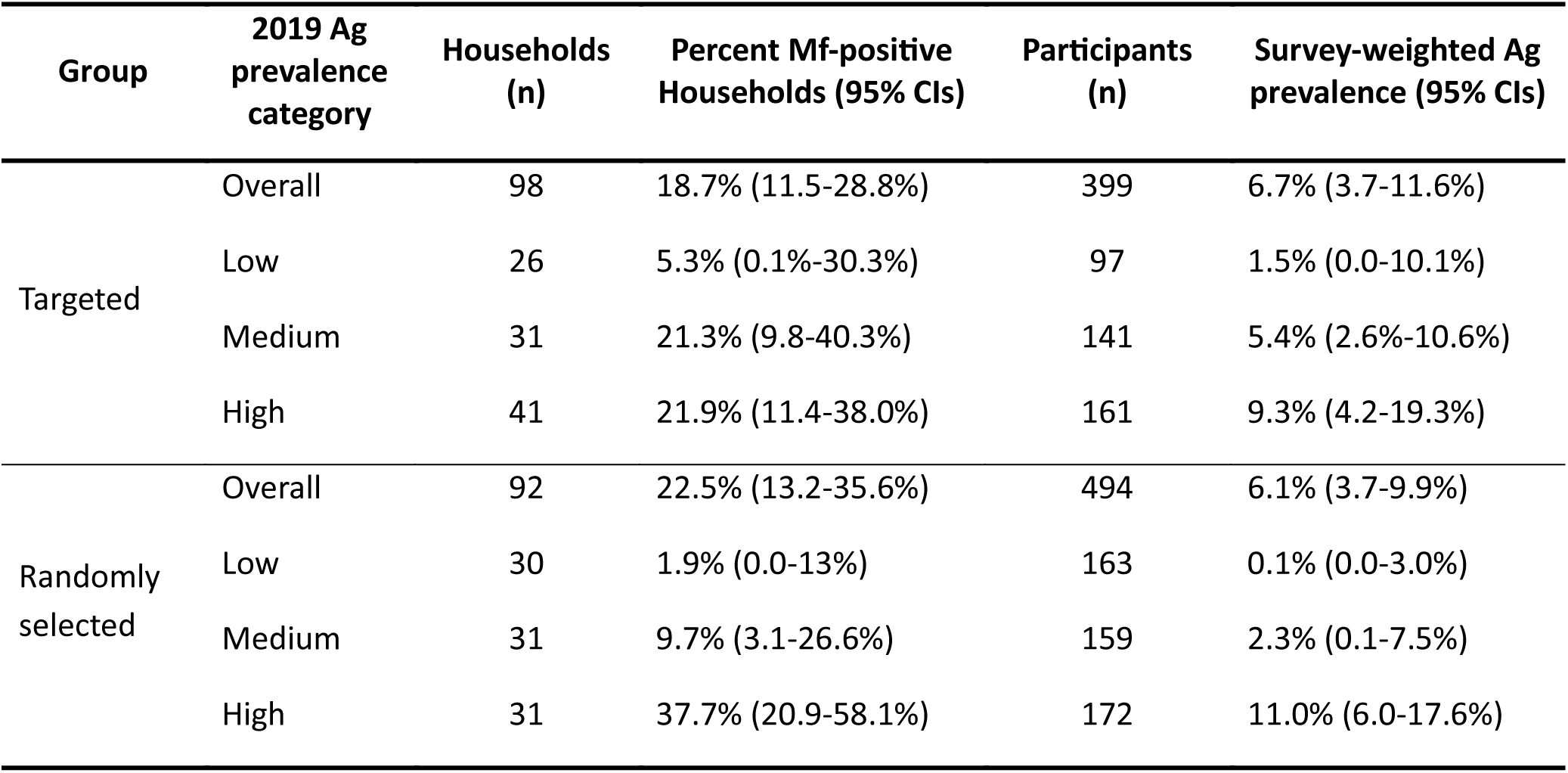
Percentage of Mf-positive households and adjusted Mf prevalence (with 95% confidence intervals) in the randomly selected and targeted groups for each 2019 Ag prevalence category - low (3-5%), medium (6-7%) and high (13-17%) - in six primary sampling units (PSUs) in Samoa in 2023.

**Supplementary S4 Table.** Ratio of antigen-positive individuals and microfilaria-positive individuals in the targeted group compared to the randomly selected group including 95% confidence intervals. Values presented for each 2019 Ag prevalence category - low (3-5%), medium (6-7%) and high (13- 17%) - in six primary sampling units (PSUs) in Samoa in 2023.

| Antigen |  | Microfilaria |  |
| --- | --- | --- | --- |
| 2019 Ag-prevalence category | Ratio (95% CI) | 2019 Ag-prevalence category | Ratio (95% CI) |
| Low | 1.12 (0.52, 3.05) | Low | 1.68 (0.30, 66.37) |
| Med | 2.38 (1.25, 5.21) | Med | 2.63 (0.90, 12.76) |
| High | 1.51 (1.02, 2.36) | High | 1.34 (0.76, 2.58) |
| Overall | 1.81 (1.32, 2.55) | Overall | 1.78 (1.10, 3.11) |

